# Hematologic complications in patients exposed to PARP inhibitors

**DOI:** 10.1101/2025.07.01.25329998

**Authors:** Joseph M. Cannova, Muriel R. Battaglia, Gregory W. Roloff, Sinan Cetin, Michael Tallarico, Wendy Stock, Anand A. Patel, Olatoyosi Odenike, Richard A. Larson, Michael J. Thirman, Mariam T. Nawas, Girish Venkataraman, Adam S. DuVall, Michael W. Drazer

**Author notes:** **Corresponding author:** Michael W. Drazer, MD, PhD, 5841 S. Maryland Avenue, MC 2115, Chicago, IL, 60637. **Data Sharing Statement:** Genomic data are available on request. **Author Contributions:** MWD and ASD conceived the study; MWD, ASD, JMC, MRB, SC, and GWR collected and analyzed the data; MWD, JMC, GWR, MT, WS, AAP, OO, RAL, MJT, MTN, GV, ASD cared for the patients; GV performed molecular pathology testing and reporting; JMC and MWD drafted the manuscript; all authors edited the manuscript.

## Abstract

Patients with *BRCA1/2-*mutated ovarian, breast, prostate, or pancreatic tumors can be treated with poly ADP-ribose polymerase (PARP) inhibitors. PARP inhibitors, however, are known to cause therapy-related myeloid neoplasms (t-MN) in a subset of patients. Predisposing factors to t-MN development in the context of PARP inhibitor exposure are not well described.

To determine the frequency of t-MN in these patients, an institutional cohort of 265 patients with exposure to PARP inhibitors was identified. A subset of these patients with PARP inhibitor-related cytopenias underwent bone marrow biopsies. Among 265 patients, 17 (6.4%) underwent a bone marrow biopsy, which yielded a therapy-related hematologic diagnosis in 47% (8/17). Breast cancer metastasis to the marrow was found in one patient, and hemophagocytic lymphohistiocytosis was found in another.

We analyzed the molecular characteristics of t-MNs in 13 PARP inhibitor-exposed patients, including five additional PARP inhibitor-exposed patients diagnosed with t-MNs in community practices. Among patients with t-MNs, five had acute therapy-related myeloid leukemia (t-AML), six were diagnosed with therapy-related myelodysplastic syndrome (t-MDS), and two had therapy-related clonal cytopenias of uncertain significance (t-CCUS). Complex karyotypes were found in four of seven patients who underwent karyotyping (57%). Next-generation sequencing identified *TP53* mutations in 7 of 9 patients analyzed (78%). Among patients with germline testing, four (40%) did not have a germline mutation identified, four (40%) had a *BRCA1* pathogenic/likely pathogenic (P/LP) variant, and two (20%) had a *BRCA2* P/LP variant. Four patients received supportive care and/or observation after blood cancer diagnosis, and six received t-MN-directed therapy. The median survival for patients who received t-MN-directed treatment was 148 days.

While cytopenias, particularly anemia, are known to occur with PARP inhibitor therapy, a subset of patients develop chronic cytopenias requiring bone marrow biopsy to evaluate for t-MN. Our study informs the expected findings of such biopsies.

## Introduction

Poly ADP-ribose polymerase (PARP) inhibitors are a treatment for solid tumors with mutations in *BRCA1, BRCA2*, or *ATM*, each of which causes homologous repair deficiency and increases dependence on alternative DNA repair mechanisms.^1^ Phase II study of olaparib in patients with germline *BRCA2-*mutations suggested responses in multiple solid tumors, with approved indications indications now including ovarian, fallopian tube, breast, prostate, pancreatic, and primary peritoneal cancers with *BRCA1/2* or homologous repair deficiency mutations.^1,2,3^

Recent studies evaluating PARP inhibitor exposure demonstrate an increased risk of reversible cytopenias, therapy-related clonal cytopenias, and therapy-related blood cancers.^4–7^ In the SOLO2 trial investigating olaparib as maintenance therapy for platinum-sensitive relapsed ovarian cancer with a *BRCA1/2* mutation, 19% of olaparib-exposed patients developed grade three anemia, and 8% of olaparib-exposed patients developed therapy-related myeloid neoplasms (t-MNs), increased from 4% in the placebo arm.^4^ Unlike t-MNs caused by exposure to alkylating agents or radiation, which may present 5 to 7 years after exposure, t-MNs secondary to PARP inhibitors may present earlier, with initial studies showing a median of 3.0 years after PARP inhibitor exposure.^4^ Subsequent pharmacovigilance studies in PARP inhibitor-treated ovarian cancer patients demonstrated therapy-related myelodysplastic syndrome and therapy-related acute myeloid leukemia may occur earlier, at a median of 211 or 355 days after exposure, respectively.^6^ The cause of the discrepancy in latency periods is unknown, although patients with ovarian malignancies are also treated with platinum agents, which may contribute to the development of hematologic neoplasms.

The high frequency of cytopenias with PARP inhibitor exposure results in referrals to hematology clinics for further evaluation. Here, we reviewed the outcomes of all patients treated with PARP inhibitors at our institution. Additionally, we analyzed the utility of bone marrow biopsies for cytopenic patients exposed to PARP inhibitors. We also assessed clinical and laboratory findings for any association with t-MN development in PARP inhibitor-exposed patients.

## Methods

After obtaining Institutional Review Board approval, we reviewed institutional records from January 2014 to November 2023. We identified 265 adult patients prescribed a PARP inhibitor. We identified 17 patients who had a bone marrow biopsy. We also reviewed our leukemia registry and identified five patients with blood cancer and prior exposure to PARP inhibitors. All diagnoses used the 2022 ICC guidelines.^8^ We used the Kaplan-Meier method to assess overall survival and Student’s T-test to compare peripheral blood counts between patients with and without a t-MN.

## Results

Of the 270 PARP inhibitor-exposed patients (265 institutional and five referred to our practice after exposure to a PARP inhibitor in community practices), 22 patients had a bone marrow biopsy performed for cytopenias after PARP inhibitor treatment (**Figure 1A**). Five of these patients were exposed to a PARP inhibitor and referred to our institution from community practices after developing a t-MN. Among the 22 patients with biopsies, thirteen (59%) were diagnosed with a t-MN, one (5%) had cytomegalovirus-driven hemophagocytic lymphohistiocytosis (HLH), and one (5%) had a myelophthisic process. Excluding the five patients referred from the community with an existing t-MN diagnosis, 6% (17/265) of the patients in our institutional cohort underwent bone marrow biopsy, and the t-MN incidence was 3.0% (8/265) among patients with confirmed PARP inhibitor exposure at our institution (**Figure 1B)**. Bone marrow biopsy resulted in a diagnosis in 59% (10/17), with the remainder attributed to therapy toxicity. A t-MN or therapy-related high-risk blood disorder (clonal cytopenia of uncertain significance, “CCUS”) was diagnosed in 47% (8/17) of patients with biopsies (**Figure 1C)**. Among patients diagnosed with a t-MN, two had therapy-related CCUS, six had therapy-related myelodysplastic syndrome, and five had therapy-related acute myeloid leukemia. The clinical and laboratory features of these patients are summarized in **Table 1** alongside those of patients who underwent a bone marrow biopsy without a t-MN diagnosis.

**Table 1.** Laboratory and clinical features of 22 PARP inhibitor-exposed patients who underwent bone marrow biopsy evaluation for a therapy-related myeloid neoplasm. WBC: white blood cell count; MCV: mean corpuscular volume. Values in parentheses show the interquartile range (IQR). Student’s T-test was used to compare the two groups.

| Characteristics at bone marrow biopsy | Patients diagnosed with blood cancer (n=13) | Patients without blood cancer (n=9) | *p* |
| --- | --- | --- | --- |
| Leukocyte count ($10^3/\mu\text{L}$) | 3.5 (2.7 - 5.3) | 5.8 (3.4 - 8.9) | 0.06 |
| Median Neutrophil Count ($10^3/\mu\text{L}$) | 1.5 (0.8 - 2.5) | 3.59 (1.3 - 6.7) | **0.02** |
| Median Hemoglobin (g/dL) | 8.7 (7.5 - 10.6) | 10.2 (7.6 - 11.4) | 0.7 |
| Median MCV (fL) | 91.3 (87.3 – 113.6) | 100.6 (99.6 – 108.1) | 0.5 |
| Median Platelets ($10^3/\mu\text{L}$) | 51 (40 - 92) | 140 (106 – 249) | **0.02** |
| Median Peripheral Blasts (%) | 0 (0 - 2) | 0 (0 - 0) | 0.1 |
| Number of Cytopenias | 2.3 (2 - 3) | 1.7 (1 - 2) | 0.17 |
| Median age at first cancer diagnosis (years) | 62 (50 - 65) | 56 (50 - 68) | 0.6 |
| Primary Tumor Type (*n*) |  |  | 0.5 |
| Ovarian Cancer (*n*, %) | 9 (69) | 8 (89%) |  |
| Breast Cancer (*n*, %) | 1 (8%) | 1 (11%) |  |
| Prostate Cancer (n, %) | 2 (15%) | 0 |  |
| Pancreatic Cancer (n, %) | 1 (8%) | 0 |  |
| Median age at bone marrow biopsy | 66 (59 - 72.5) | 58 (56 - 68) | 0.65 |
| Known Germline Mutation (n, %) | 8 (61.5%) | 4 (44.4%) | 0.4 |
| PARP inhibitor |  |  | 0.4 |
| Olaparib (n, %) | 10 (83%) | 6 (60%) |  |
| Rucaparib (n, %) | 1 (8%) | 3 (30%) |  |
| Niraparib (n, %) | 1 (8%) | 1 (10%) |  |
| Median duration of PARP inhibitor exposure (Days, IQR) | 506 (231 - 817) | 250 (213 - 639) | 0.15 |

**Figure 1.**
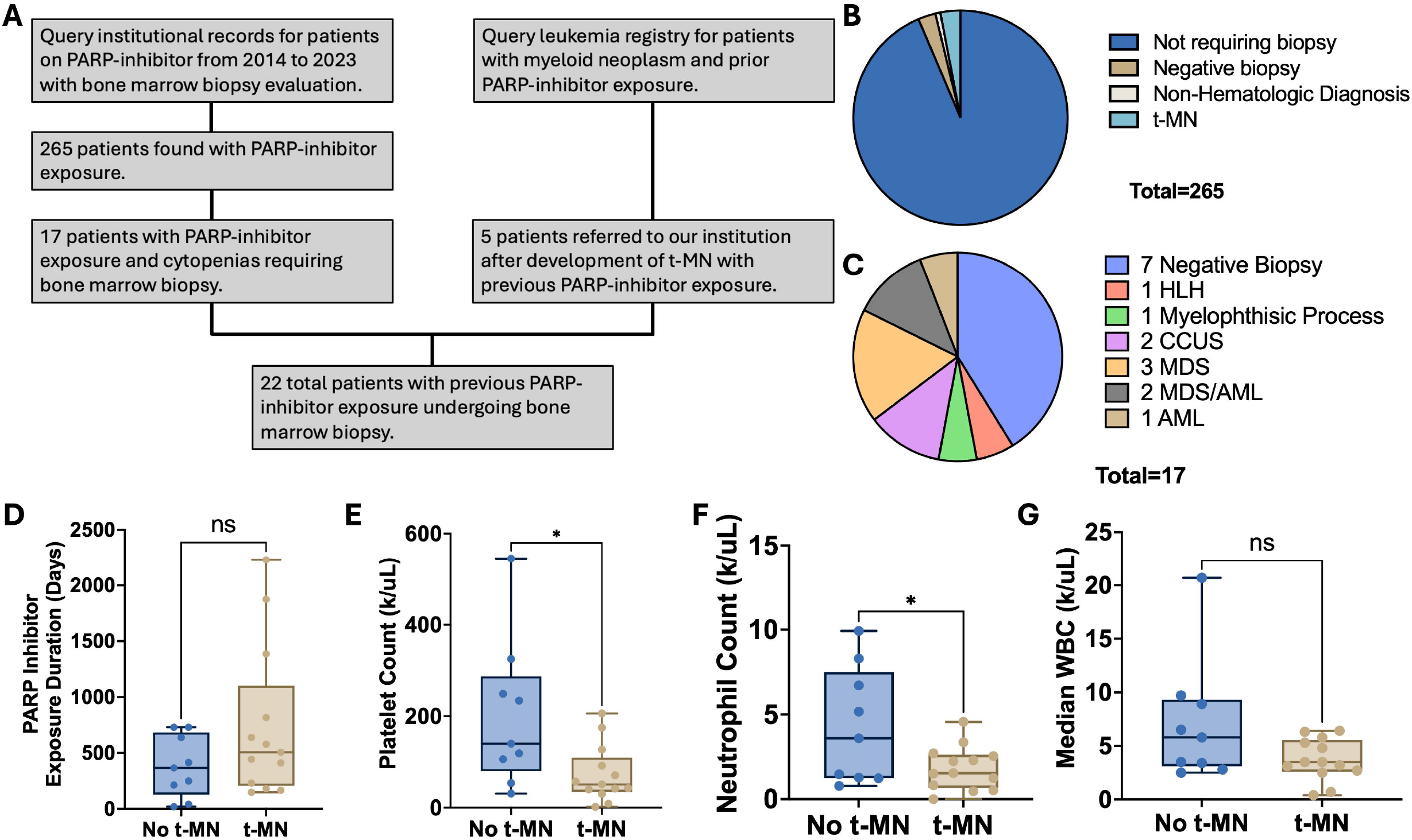
**A**. CONSORT Plot of patients included in this study. **B**. Pie plot showing the proportion of patients who did not undergo bone marrow biopsies (n=248) and patients who underwent bone marrow biopsies (n=17), those with non-hematologic findings (n=2), negative biopsy (n=7), and t-MN (n=8) **C**. Pie plot showing diagnoses from bone marrow biopsies: no diagnosis (n=7), HLH (n=1), myelophthisic process (n=1), t-CCUS (n=2), t-MDS (n=3), t-MDS/AML (n=2), and t-AML (n=1). ∫D. Duration of PARP inhibitor exposure in patients with (gold) and without (blue) diagnoses of therapy-related myeloid neoplasms (t-MN). **E**. Platelet count in patients with (gold) and without (blue) diagnoses of t-MN. **F**. Neutrophil count in patients with (gold) and without (blue) diagnoses of t-MN. **G**. White blood cell count in patients with (gold) and without (blue) diagnoses of t-MN. * denotes p<0.05

We then determined if laboratory and clinical parameters differed between the 13 patients diagnosed with a t-MN and the nine patients with a biopsy who were not diagnosed with a t-MN. Compared to the patients without t-MN diagnoses, leukocytes were not reduced (p=0.06), but neutrophils (p=0.03) and platelets (p=0.02) were lower in the t-MN group (**Table 1, Figure 1C-1F**). Despite t-MN diagnoses, most patients did not have detectable peripheral blasts.

The median survival for patients with high-risk t-MN was 159 days (n=9) and 148 days for patients receiving treatment (n=8, **Figure 2A and 2B**). These patients were treated with hypomethylating agents in combination with venetoclax or on clinical trials. None received induction with intensive chemotherapy or allogeneic stem cell transplant.

**Figure 2.**
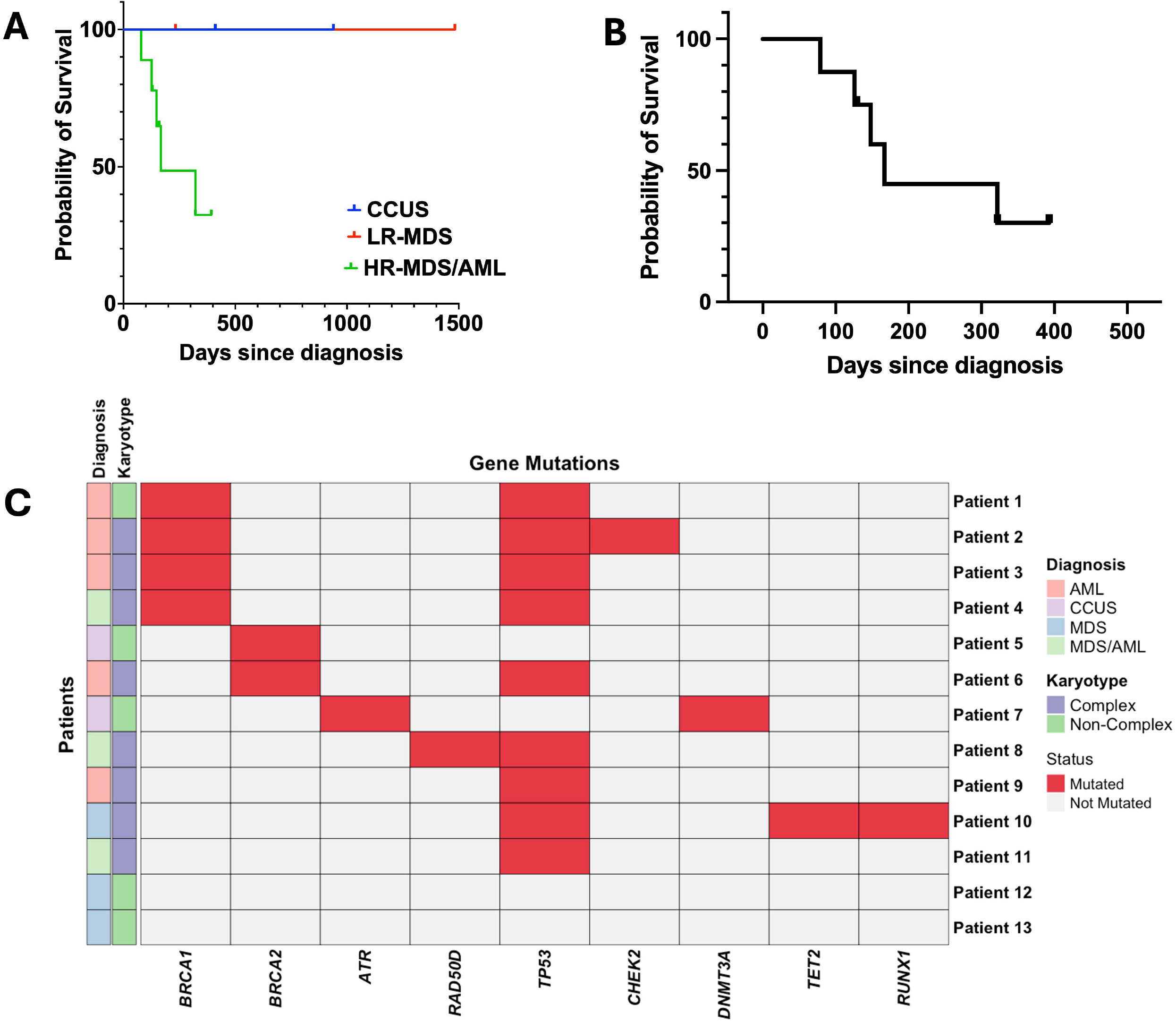
**A**. Survival curve of patients diagnosed with CCUS (mOS NR, n=2), low-risk myelodysplastic MDS (n=2 mOS NR), and high-risk MDS or AML (mOS 148 days, n=9). Deaths are censored if unrelated to t-MN diagnosis. **B**. Survival curve of patients with HR-MDS/AML who received treatment (mOS 148 days, n=8). **C**. Co-mutation plot for patients who developed therapy-related blood disorders. Patients were divided into CCUS: clonal cytopenia of uncertain clinical significance; LR-MDS: low risk myelodysplastic syndrome; HR-MDS: high-risk MDS; and AML: acute myeloid leukemia; mOS: median overall survival.

We analyzed other factors, including age at biopsy, number of cytopenias, duration of PARP inhibitor exposure, type of PARP inhibitor, solid tumor diagnosis, and the presence of germline mutations (**Table 1**). These variables did not differ significantly between patients with and without t-MNs. The t-MNs that developed in PARP-exposed patients were enriched for *TP53* mutations (n=9/13, 69%), as has been reported by others (**Figure 2C)**.^5, 9, 10^ Of these patients, four (44% of *TP53*-mutated and 31% of the total t-MN cohort) had biallelic *TP53* mutations, an adverse risk factor.^11^ Similarly, among eleven patients with available data, eight (77%) had a complex or adverse-risk karyotype, consistent with other studies.^9, 10^

## Discussion

This study provides insights into the genomic risk factors for developing t-MN. We hypothesized that germline mutations in *BRCA1, BRCA2*, or other DNA damage response genes would result in increased risk of t-MN development in patients with germline mutations (**Figure 2B**). However, no significant difference in the distribution of germline mutations was observed across patients with t-MN and those without t-MN (**Table 1**). However, this signal may be reduced by the relatively small number of patients with t-MNs in our cohort. The diagnosis of t-MNs in patients without germline mutations suggests that PARP inhibitors may increase t-MN risk in patients regardless of germline mutational status.

Additionally, based on reports that PARP inhibitors increase the risk of clonal hematopoiesis (CH), including CH that is partially reversible after stopping PARP inhibitors, we hypothesized that our cohort would be enriched in patients with CH or clonal cytopenias. However, we found CH-associated mutations in only three patients with bone marrow biopsies (18%). The bone marrow sample from one patient with a germline *BRCA2* (p.K1381fs*) mutation had a clonal del(7q) (3.5% of cells by FISH) and no additional molecular mutations on NGS. A classic CH-associated *DNMT3A* mutation (p.Q816* variant allele frequency, 7%) was detected in a patient with thrombocytopenia and normal bone marrow findings. The CH-related *TET2* mutation (p.S271fs, variant allele frequency 31%) was found in a third patient diagnosed with a therapy-related myelodysplastic syndrome while taking rucaparib. Though this study did identify t-CCUS patients, no patients diagnosed with t-MN in this cohort demonstrated molecular or laboratory evidence of antecedent t-CCUS (**Figure 2B**).^15^

Notably, the t-MNs in our patient cohort had a relatively low peripheral blast burden despite adverse-risk molecular characteristics, making bone marrow evaluation essential for t-MN diagnosis in these patients. For patients on PARP inhibitors at the time of t-MN diagnosis, peripheral blasts ranged from 0 to 5%, with a higher bone marrow blast percentage (0-70%). Intriguingly, no difference in duration of PARP inhibitor exposure was found between patients with t-MNs and those without (p=0.3). We noted that one patient with overt dysplasia and therapy-related myelodysplastic syndrome had morphologic improvement in their bone marrow and disappearance of a cytogenetically abnormal clone after discontinuing the PARP inhibitor. A similar pattern has been reported with PARP inhibitor-associated CH, which can regress with drug discontinuation.^13^

## Conclusion

In conclusion, this study demonstrates the diagnostic importance of pursuing bone marrow biopsy in PARP inhibitor-exposed patients with persistent leukopenia, neutropenia, or thrombocytopenia. We found that 6.4% of patients in one of the largest single-institution series of patients exposed to PARP inhibitors ultimately had bone marrow biopsies performed. Among our PARP-exposed patients, 3.0% developed therapy-related blood cancers or high-risk blood disorders, including myelodysplastic syndrome, acute myeloid leukemia, and therapy-related CCUS. These patients were enriched with high-risk molecular features, including *TP53* mutations and complex karyotypes. The median survival for our patients with t-MNs was 148 days, which reflects the aggressive biology of these diseases in PARP-exposed patients. We did not observe molecular evidence of antecedent CH or CCUS in most patients in this cohort; however, the small sample size of patients with bone marrow biopsies limited our analysis. This cohort demonstrates the frequent diagnosis of t-MN despite low or absent peripheral blasts, supporting the use of bone marrow biopsy in patients with PARP inhibitor exposure and chronic, unexplained cytopenias. T-MNs occurred in patients with and without germline mutations, so blood cancer risk in PARP-exposed patients is increased regardless of the presence of germline mutations. Our findings suggest that further research is needed into the mechanisms driving PARP inhibitor-associated hematologic toxicity, and that PARP inhibitors should only be used in clinical settings where clear survival benefits have been demonstrated.

## Data Availability

Genomic data are available on request

## ACKNOWLEDGMENTS

We thank our patients who participated in this research. This work was supported by the National Institutes of Health Clinical Therapeutics award (JMC) and the National Institutes of Health Loan Repayment Program.

## Bibliography & References Cited

1. Drew Y, Zenke FT, Curtin NJ. DNA damage response inhibitors in cancer therapy: lessons from the past, current status and future implications. Nat Rev Drug Discov. 2025;24:19–39.

2. Kaufman B, Shapira-Frommer R, Schmutzler RK, et al. Olaparib monotherapy in patients with advanced cancer and a germline BRCA1/2 mutation. J Clin Oncol. 2015;33:244–250.

3. Mateo J, Carreira S, Sandhu S, et al. DNA-Repair Defects and Olaparib in Metastatic Prostate Cancer. N Engl J Med. 2015;373:1697–1708.

4. Poveda A, Floquet A, Ledermann JA, et al. Olaparib tablets as maintenance therapy in patients with platinum-sensitive relapsed ovarian cancer and a BRCA1/2 mutation (SOLO2/ENGOT-Ov21): a final analysis of a double-blind, randomised, placebo-controlled, phase 3 trial. Lancet Oncol. 2021;22:620–631.

5. Marmouset V, Decroocq J, Garciaz S, et al. Therapy-related Myeloid Neoplasms Following PARP Inhibitors: Real-life Experience. Clin Cancer Res. 2022;28:5211–5220.

6. Zhao Q, Ma P, Fu P, et al. Myelodysplastic Syndrome/Acute Myeloid Leukemia Following the Use of Poly-ADP Ribose Polymerase (PARP) Inhibitors: A Real-World Analysis of Postmarketing Surveillance Data. Front Pharmacol. 2022;13:912256.

7. Morice PM, Leary A, Dolladille C, et al. Myelodysplastic syndrome and acute myeloid leukaemia in patients treated with PARP inhibitors: a safety meta-analysis of randomised controlled trials and a retrospective study of the WHO pharmacovigilance database. Lancet Haematol. 2021;8:e122–e134.

8. Arber DA, Orazi A, Hasserjian RP, et al. International Consensus Classification of Myeloid Neoplasms and Acute Leukemias: integrating morphologic, clinical, and genomic data. Blood. 2022;140:1200–1228.

9. Oliveira JL, Greipp PT, Rangan A, Jatoi A, Nguyen PL. Myeloid malignancies in cancer patients treated with poly(ADP-ribose) polymerase (PARP) inhibitors: a case series. Blood Cancer J. 2022;12:11.

10. Todisco E, Gigli F, Ronchini C, et al. Hematological disorders after salvage PARP inhibitor treatment for ovarian cancer: Cytogenetic and molecular defects and clinical outcomes. Int J Cancer. 2022;151:1791–1803.

11. Nawas MT, Kosuri S. Utility or futility? A contemporary approach to allogeneic hematopoietic cell transplantation for TP53-mutated MDS/AML. Blood Adv. 2024;8:553–561.

12. Valenza C, Mongillo M, Gigli F, et al. Germline BRCA pathogenic variants in patients with ovarian cancer and post-poly (ADP-ribose) polymerase inhibitor myeloid neoplasms. ESMO Open. 2024;9:103685.

13. Nuttall Musson E, Miller RE, Mansour MR, Lockley M, Ledermann JA, Payne EM. Monitoring clone dynamics and reversibility in clonal haematopoiesis and myelodysplastic neoplasm associated with PARP inhibitor therapy-a role for early monitoring and intervention. Leukemia. 2024;38:215–218.

14. Marshall CH, Gondek LP, Daniels V, et al. Association of PARP inhibitor treatment on the prevalence and progression of clonal hematopoiesis in patients with advanced prostate cancer. Prostate. 2024;84:954–958.

15. Jaiswal S, Fontanillas P, Flannick J, et al. Age-related clonal hematopoiesis associated with adverse outcomes. N Engl J Med. 2014;371:2488–2498.

